# Omicron-specific cytotoxic T-cell responses are boosted following a third dose of mRNA COVID-19 vaccine in anti-CD20-treated multiple sclerosis patients

**DOI:** 10.1101/2021.12.20.21268128

**Authors:** Natacha Madelon, Nelli Heikkilä, Irène Sabater Royo, Paola Fontannaz, Gautier Breville, Kim Lauper, Rachel Goldstein, Alba Grifoni, Alessandro Sette, Claire-Anne Siegrist, Axel Finckh, Patrice H. Lalive, Arnaud M. Didierlaurent, Christiane S. Eberhardt

**Affiliations:** Center for Vaccinology, Department of Pathology and Immunology, Faculty of Medicine, University of Geneva, Geneva, Switzerland; Department of Neurosciences, Division of Neurology, University Hospital of Geneva & Faculty of Medicine, University of Geneva, Geneva, Switzerland; Department of Medicine, Division of Rheumatology, University Hospital of Geneva & Faculty of Medicine, University of Geneva, Geneva, Switzerland; Center for Infectious Disease and Vaccine Research, La Jolla Institute for Immunology, University of California, San Diego, La Jolla, USA; Department of Medicine, Division of Infectious Diseases and Global Public Health, University of California, San Diego, La Jolla, USA; Division of General Pediatrics, Department of Woman, Child and Adolescent Medicine, Faculty of Medicine, University of Geneva; and Center for Vaccinology, Geneva University Hospitals, Geneva, Switzerland; Emory Vaccine Center, Emory University School of Medicine, Atlanta, USA

**Keywords:** Cytotoxic T-cell responses, SARS-CoV-2, Omicron, Delta, anti-CD20, mRNA vaccination

## Abstract

**Importance:** The SARS-CoV-2 variant Omicron escapes neutralizing antibody responses elicited after COVID-19 vaccination, while T-cell responses might be better conserved. It is crucial to assess how a third dose of vaccination modifies these responses, particularly for immunocompromised patients with readily impaired antibody responses.

**Objective:** To determine T-cell responses to the Spike (S)-protein of Omicron in anti-CD20 treated patients before and after their third mRNA COVID-19 vaccination

**Design:** Prospective observational monocentric study

**Setting:** Conducted since March 2021 at the University Hospital Geneva

**Participants:** Twenty adults with multiple sclerosis on anti-CD20 treatment (ocrelizumab) who received their third dose of mRNA COVID-19 vaccine 6 to 7 months after their second vaccination. Intervention: Blood sampling before and one month after the third vaccine dose

**Main outcomes and measures:** Quantification of CD4 and CD8 (cytotoxic) T cells specific for SARS-CoV-2 S-protein of vaccine strain, Delta and Omicron variants, using activation marker induced assay (AIM) and comparing frequencies before and after the third vaccine dose.

**Results:** S-specific CD4 and CD8 T-cell memory against all variants was maintained in around half of the patients six months after their second vaccination, albeit at lower frequencies against Delta and Omicron variants. A third dose enhanced the number of responders to all variants and significantly increased CD8 T-cell responses. The frequencies of T cells specific to Omicron and Delta remained lower than those specific to the vaccine strain after the boost.

**Conclusion and relevance:** Vaccinated MS patients on anti-CD20 treatment show robust T-cell responses that recognize S from the circulating Delta and Omicron variants. Response rates increased after the third dose, demonstrating that a booster dose might improve cytotoxic T-cell mediated protection against severe disease in patients with low humoral response. The clinical relevance of the reduced frequencies of T cells specific to Omicron will need to be monitored in the future.

**Key points:** *Question:* Are T-cell responses to Omicron variant conserved in anti-CD20 treated MS patients after COVID-19 mRNA vaccination?

*Findings:* Omicron Spike-specific CD4 and CD8 T cells were detectable in around half of twenty patients six months after the second COVID-19 vaccine dose, and cytotoxic T-cell responses increased following the third dose. Frequencies of T cells specific against the S-protein of Delta and Omicron were lower compared to the vaccine strain, both before and after boost.

*Meaning:* In anti-CD20-treated MS patients the vaccine-induced T-cell responses are little affected by the mutations carried by Omicron, and a third vaccine dose improves cytotoxic T-cell responses.

## Introduction

The SARS-CoV-2 variant Omicron was designated a variant of concern (VOC) on November 26, 2021^1^ and is spreading rapidly. The protection provided by current COVID-19 vaccines is not established. Preliminary studies suggest that antibodies neutralizing Omicron, which carries multiple mutations in the spike protein (S_Omicron_), critically wane in the months following heterologous infection or vaccination^2,3^. A third vaccine dose seems to boost neutralizing antibodies to, but less efficiently than for the Delta variant^2^. Whether the mutations in S_Omicron_ impact T-cell recognition, which is known to be less affected than antibody responses for other variants^4^, is still under investigation. SARS-CoV-2-specific T-cell responses have a role in protection against severe COVID-19 and their importance is further underlined when antibody responses to vaccination are defective. In individuals with multiple sclerosis (MS) on anti-CD20 treatment, COVID-19 mRNA vaccines induce suboptimal antibody responses but robust and functional T-cell responses^5^. In the current situation and specifically for this vulnerable population, it is crucial to understand if Omicron-specific T cells are elicited by two doses of mRNA vaccine and potentially increased by a third dose. This is of critical clinical relevance since some therapeutic monoclonal anti-SARS-CoV-2 antibodies that are given to these patients to alleviate symptoms are reported to be ineffective against Omicron^3^. In the present study, we report the T-cell responses to Omicron compared to vaccine and Delta variants in anti-CD20 treated patients, before and following the third dose of COVID-19 mRNA vaccines.

## Methods

This prospective observational study has been carried out since March 2021 at the Geneva University Hospitals, Switzerland, according to the principles of Good Clinical Practice and was approved by the Geneva Cantonal Ethics Commission (2021-00430). Informed consent was obtained from all participants and more study details are found elsewhere^5^. For the herein performed analyses, adult individuals who were on anti-CD20 treatment (ocrelizumab) for MS and who received their third dose of mRNA COVID-19 vaccine before November 1st, 2021 were included. Serum and peripheral blood mononuclear cells (PBMC) were collected on the day of vaccination and 30 days after the third dose of COVID-19 vaccine. Anti-SARS-CoV-2 nucleoprotein (anti-N) total antibodies and anti-SARS-CoV-2 RBD (anti-receptor binding domain) total antibodies were measured, and seroconversion was defined as > 0.8 IU/ml for the anti-RBD antibodies and > 1 cut-off index (COI) for anti-N antibodies as described previously^5^.

For the activation induced marker (AIM) assay, PBMC were stimulated overnight with 1µg/ml of SARS-CoV-2 spike peptide megapools (overlapping 15mers by 10 amino acids spanning the entire antigen of vaccine, Delta, and Omicron strains), DMSO as negative and SEB as positive control. Cells were stained with fluorescent-labeled antibodies and analyzed by flow cytometry using BD LSR Fortessa. Statistical analysis was performed with GraphPad Prism software (version 8.0.2). Quantitative variables were compared by Wilcoxon signed rank test and p values < 0.05 was considered statistically significant.

## Results

We included 20 patients with MS under anti-CD20 monotherapy (Ocrelizumab), who received a third dose of COVID-19 mRNA vaccine, either mRNA-1273 (100µg, n=16) or BNT162b2 (n=4). The median interval between the second and the third vaccine dose was 26.7 weeks (IQR 22.3-29.0). Before the booster dose, 55% of patients were seropositive for anti-RBD-antibodies, increasing to 65% one month after the third dose. One patient had been infected before vaccination as witnessed by detectable anti-N antibodies (Demographic details Table 1).

**Table 1.** Demographic details and serological results

|  | **MS patients** |
| --- | --- |
| **n** | 20 |
| **Female, n(%)** | 9 (45) |
| **Age, median[IQR]** | 46 [39.3, 59.9] |
| **Anti-CD20 therapy, n(%)<sup>1</sup>** |  |
| **Ocrelizumab (600 mg)** | 20 (100) |
| **History COVID-19 (RT-PCR), n (%)** | 1 (5) |
| **Positive anti-N serology before the 3<sup>rd</sup> dose, n(%)** | 1 (5) |
| **Vaccine, n(%)** |  |
| **BNT162b2** | 4 (20) |
| **mRNA-1273** | 16 (80) |
| **Interval between 2nd and 3rd dose weeks [IQR]** | 26.7 [22.3-29.0 ] |
| **Anti-RBD Ig antibody GMC U/ml [SD] at D30 post-3rd dose** | 52.8 [72.9] |
| **Seropositive (> 0.8U/ml) at D30 post-3rd dose n (%)** | 13 (65) |

<sup>1</sup>The dose mentioned is the total dose that the individual received in around 2 weeks.

AIM+ cytotoxic (CD8+) T cells specific for the S_Vaccine_, S_Delta_, and S_Omicron_ protein were present before the booster dose in respectively 60%, 50% and 45% of patients. Following the booster dose, the fraction of responders increased from 75% for S_Vaccine_ to 70% for both S_Delta_ and S_Omicron_, but the differences remained statistically non-significant for all strains. Nevertheless, the frequency of AIM+ S-specific CD8 T cells was clearly enhanced after the third dose for all variants (Figure 1A+B). S-specific AIM+ CD8 T cells had mostly an effector memory phenotype which was further increased after the booster dose, again irrespective of the variant (Figure 1C). However, the frequencies of both S_Delta_ and S_Omicron_-specific AIM+ CD8 T cells in responders were significantly lower than the ones specific to S_Vaccine_ (Figure 1D). Before the third dose, the median frequency of S_Delta_ and S_Omicron_ - responses was 83.0% (95%CI 73.6-115%) and 79.0% (95%CI 59.4-100%) that of the S_Vaccine_-responses, respectively. After boosting, the frequency of variant-specific CD8 T cells remained lower than those specific for the vaccine strain (89.3% (95%CI 57.6-100%) and 71.0% (95%CI 41.6-96.2%), respectively for S_Delta_ and S_Omicron_). No difference was observed between the levels of CD8 T cells specific to Delta and Omicron (Suppl. Figure 1 C).

**Figure 1:**
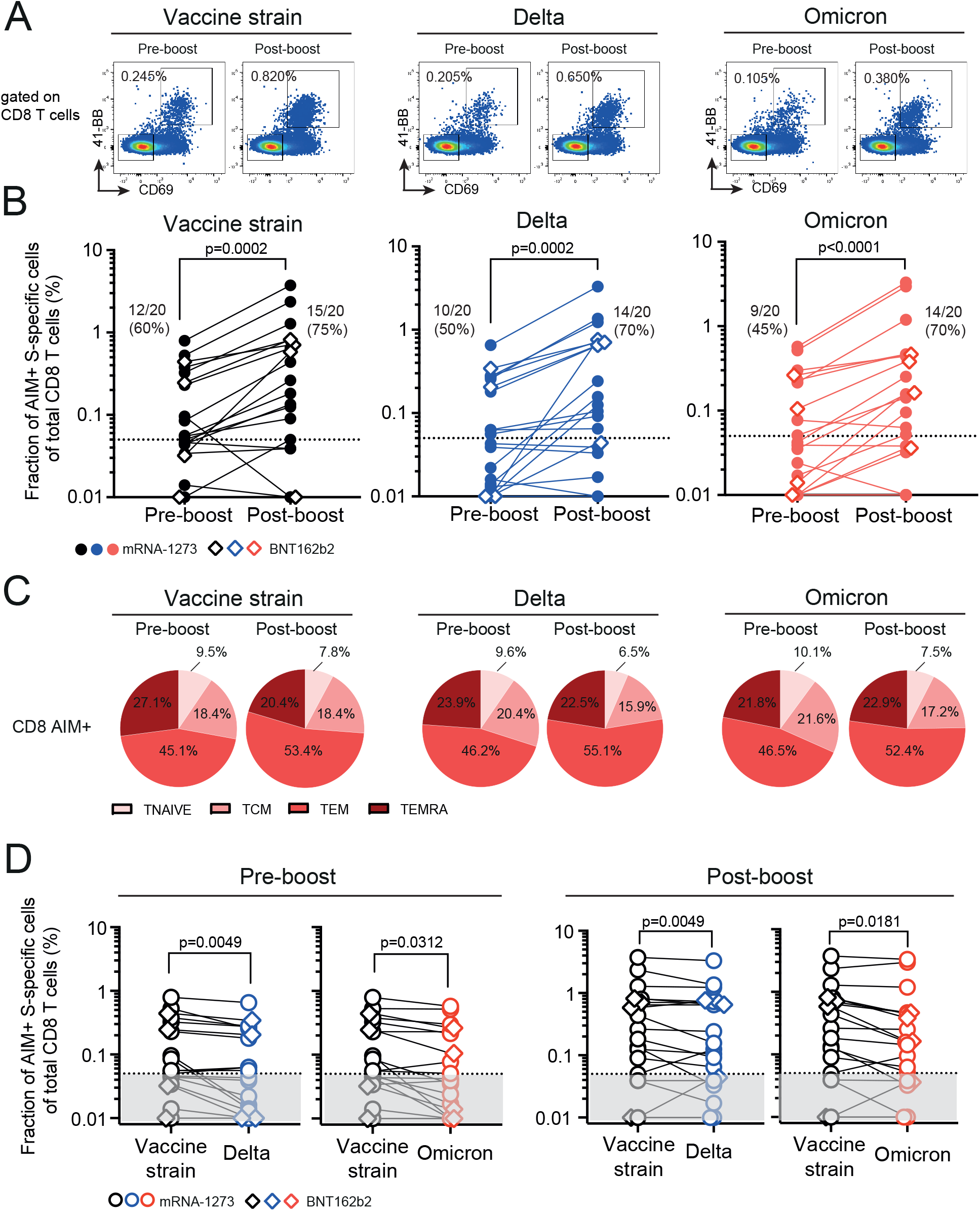
CD8 T-cell responses specific to vaccine strain and variants Delta and Omicron are boosted after the third vaccine dose. (A) Representative flow cytometry plots of CD8 T cells after PBMC stimulation with S-peptide pools from vaccine strain, Delta and Omicron, before and 30 days after the third vaccination. S-specific AIM+ CD8 T cells are gated as CD69+ 41BB+ cells. (B) Graphs summarize data from n=20 ocrelizumab-treated individual patients with their geometric mean. One dot represents one patient and lines connect pre-and post-boost vaccination frequencies of AIM+ CD8 T cells specific for SARS-CoV-2 vaccine strain (left), the variant Delta (middle) and Omicron (right) and are background-subtracted. The dotted line represents the limit of detection. Percentages of responders (those with level above limit of detection) are indicated. (C) Pie charts showing the proportions of memory phenotype of Spike-specific CD8 T cells of individuals with detectable AIM+ CD8 T cells for SARS-CoV-2 vaccine strain (left, n=12 pre-boost, 15 post-boost), and variants Delta (middle, n=10 pre-boost, n=14 post boost) and Omicron (right, n=9 pre-boost, n=14 post-boost). The proportions of naïve (T_NAIVE_, CD45RA^+^ CCR7^+^), central memory (T_CM_, CD45RA^-^ CCR7^+^), effector memory (T_EM_, CD45RA^-^ CCR7^-^) and RA-positive effector memory T cells (T_EMRA_, CD45RA^+^ CCR7^-^) are shown. Results of bulk non-specific CD8 T cells (AIM-CD8 T cells) are found in Suppl figure 1B. (D-E) Graphs illustrating differences in frequencies of CD8 T cells specific for (D) S_Vaccine_ and S_Delta_ and (D) S_Vaccine_ and S_Omicron_ in patients before (left) and 30 days after (right) the third vaccine dose. Wilcoxon signed rank test was used on responders to S_Vaccine_ to assess differences between variants.

The frequency of patients with detectable CD4 T-cell responses were 45-50% before the third dose, increasing to 70-75% for S_Vaccine_ and S_Delta_ but remaining at modest 55% for S_Omicron_ post-boost. Unlike cytotoxic T cells, no significant changes in the frequencies of S-specific AIM+ CD4 T cells were detected after the third dose (Figure 2A+B). The responding cells presented principally effector memory and central memory phenotypes with the frequency of the former increasing at the expense of the latter after the booster dose (Figure 2C).

**Figure 2:**
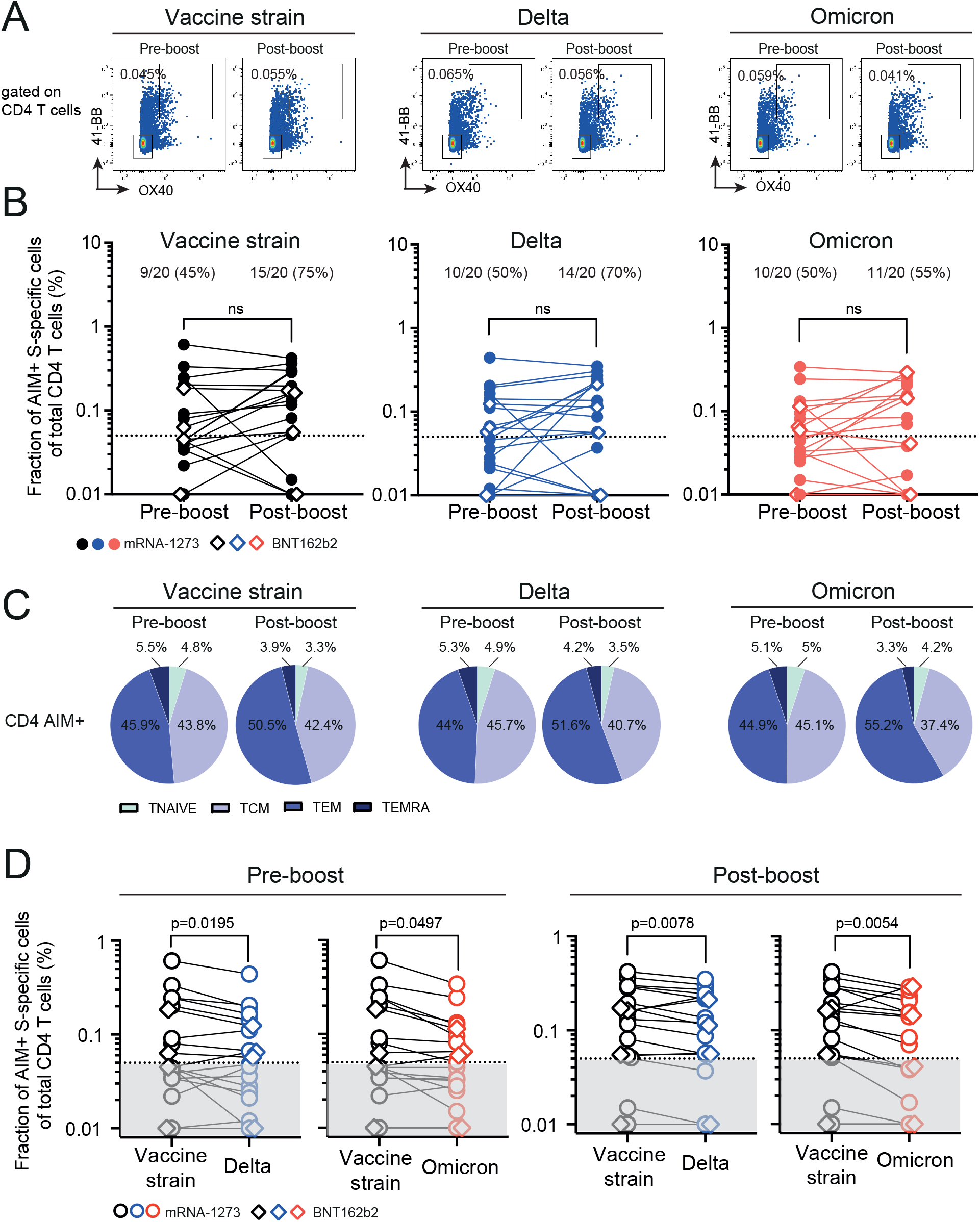
CD4 T-cell frequencies specific to vaccine strain are higher compared to the variants Delta and Omicron. (A) Representative flow cytometry plots of CD4 T cells after PBMC stimulation with S-peptide pools from the vaccine strain, Delta and Omicron, before and 30 days after the third vaccination. S-specific AIM+ CD4 T cells are gated as OX40+ 41-BB+. (B) Graphs summarize data from ocrelizumab-treated individual patients (n=20) with geometric mean. One dot represents one patient and lines connect pre- and post-boost vaccination frequencies of CD4 T cells specific for SARS-CoV-2 vaccine strain (left), variants Delta (middle) and Omicron (right) and are background-subtracted. The dotted line represents the limit of detection. Percentages of responders (those with level above limit of detection) are indicated. Patients vaccinated with mRNA-1273 are depicted as closed circles and those with BNT162b2 as open rectangles. The comparison between pre- and post-booster vaccination was calculated using with Wilcoxon signed rank test. (C) Pie charts showing the proportions of memory phenotype Spike-specific AIM+ CD4 T cells of individuals with detectable AIM+ CD4 T cells for SARS-CoV-2 vaccine strain (left, n=9 pre-boost, n=15 post boost), and variants Delta (middle, n=10 pre-boost, n=14 post boost) and Omicron (right, n=10 pre-boost, n=11 post-boost). The proportions of naïve (T_NAIVE_, CD45RA^+^ CCR7^+^), central memory (T_CM_, CD45RA^-^ CCR7^+^) and effector memory (T_EM_, CD45RA^-^ CCR7^-^) are shown. Results of bulk non-specific CD4 T cells (AIM-CD4 T cells) are found in Supplemental Figure 1A. (D) Graphs showing differences in frequencies of CD4 T cells specific for S_Vaccine_ and S_Delta_ and S_Vaccine_ and S_Omicron_ in patients before (left) and 30 days after (right) the third vaccine dose. Wilcoxon signed ranks test was used on responders to S_Vaccine_.

Similar to what was observed for CD8 T cells, S_Delta_ and S_Omicron_-specific CD4 T-cell responses were significantly reduced when compared S_Vaccine_-responses. The median frequencies after the boost were for S_Delta_ 83.5% (95%CI 69.4-105.6%) and for S_Omicron_ 72.3% (95%CI 53.4-82.7%) that of the S_Vaccine_-responses, as compared to respectively 72.2% (95%CI 67.4-90.5%) and 62.5% (95%CI 51.0-89.0%), before the boost.

## Discussion

In anti-CD20 treated patients, SARS-CoV-2-specific antibody titers are low or undetectable after two doses of mRNA vaccines and are not restored after a third dose as shown here and for lymphoma patients^5,8^. Nevertheless, T-cell responses after the second dose develop in MS patients with anti-CD20 treatment similarly to immunocompetent individuals^5,6^. With the emergence of variants such as Omicron that can evade spike-specific antibody responses, maintenance of robust vaccine-induced T-cell responses against those VOCs may offer some level of protection to vaccinees with low circulating antibodies.

Here we show that six months after the second vaccine dose, memory T-cell responses to Delta and Omicron variants are detectable in around half of the patients in line with the durability of vaccine-specific T-cell responses reported in healthy individuals^9^. The number of responders increased for both CD4 and CD8 T cells after the third dose, showing that those with undetectable memory response after the primary vaccination series can be boosted. Importantly, a third dose particularly increased the frequency of cytotoxic CD8 T cells that are important in viral defense irrespective of the SARS-CoV-2 variants. Yet, both before and after the booster dose, the frequencies of S-specific T cells were lower against Omicron. If confirmed in larger cohort, this could indicate that the recognition of binding affinities of T-cell epitopes may be compromised as mutations accumulates in VOC, as observed with Omicron variant. That being said, T-cell responses remained robust in those patients and a third dose is required to further strengthen this response. It is unknown whether the magnitude of T cells correlates with protection from severe disease, but responder rates were similar for vaccine-strain and variants.

The interpretation of our results is limited by the low sample size and the limited post-vaccination follow-up. The monitoring of breakthrough infections in this population will be important to confirm the benefit of vaccination, in particular against Omicron.

In conclusion, vaccinated MS patients on anti-CD20 treatment show robust T-cell responses that recognize S from vaccine strain, Delta and Omicron variants and increased cytotoxic T cell responses after boost. However, reduced frequencies of T cells specific to Delta and Omicron compared to vaccine strain were measured and clinical implications of these findings need to be assessed.

## Data Availability

All data produced in the present study are available upon reasonable request to the authors

## FUNDING

This work was supported by the HUG private Foundation (CSE, CAS, AD) and by the Giorgi-Cavaglieri foundation (AD).This work was additionally funded in whole or in part with Federal funds from the National Institute of Allergy and Infectious Diseases, National Institutes of Health, Department of Health and Human Services, under Contract No. 75N93021C00016 and 75N9301900065 to A.S.

## CONFLICT OF INTEREST

A.S. is a consultant for Gritstone Bio, Flow Pharma, Arcturus Therapeutics, ImmunoScape, CellCarta, Avalia, Moderna, Fortress and Repertoire. LJI has filed for patent protection for various aspects of T cell epitope and vaccine design work.

All of the other authors declare no competing interests.

## ACKNOWLEDGMENTS

We are grateful to all volunteers for their participation in the study. We thank Mélanie Jaquet, Sandra Fernandes Oliveira, Olivia Studer, Sandrine Bastard and colleagues at the Clinical Research Center, University Hospital and Faculty of Medicine, Geneva for their involvement in patient recruitment and sample collection; Chantal Tougne, Yves Donati, Wafae Adouan and Sophie Coudurier for their contributions to processing the clinical samples and support in experimental work, and Myriam Ventura-Lehnis for her administrative support. We thank Jean-Pierre Aubry-Lachainaye, Cécile Gameiro and Grégory Schneiter from the flow cytometry core facility for their technical support. We thank all colleagues that have provided support to participant recruitment, sample collection and experimental work.

---

**Supplemental figure 1: Memory phenotype of AIM-CD4 and CD8 cells and comparison of frequencies of Delta and Omicron-specific T cells**

Pie charts referring to Figure 1C and Figure 2C, showing the proportions of memory phenotype of bulk non-S-protein-specific and AIM-(A) CD8 and (B) CD4 T cells in individuals with (A) detectable AIM+ CD8 T cells for SARS-CoV-2 vaccine strain (left, n=12 pre-boost, 15 post-boost), and variants Delta (middle, n=10 pre-boost, n=14 post boost) and Omicron (right, n=9 pre-boost, n=14 post-boost). The proportions of naïve (T_NAIVE_, CD45RA^+^ CCR7^+^), central memory (T_CM_, CD45RA^-^ CCR7^+^), effector memory (T_EM_, CD45RA^-^ CCR7^-^) and RA-positive effector memory T cells (T_EMRA_, CD45RA^+^ CCR7^-^) are indicated. (B) Results of individuals with detectable AIM+ CD4 T cells for SARS-CoV-2 vaccine strain (left, n=9 pre-boost, n=15 post boost), and variants Delta (middle, n=10 pre-boost, n=14 post boost) and Omicron (right, n=10 pre-boost, n=11 post-boost). The proportions of naïve (T_NAIVE_, CD45RA^+^ CCR7^+^), central memory (T_CM_, CD45RA^-^ CCR7^+^) and effector memory (T_EM_, CD45RA^-^ CCR7^-^) and RA-positive effector memory T cells (T_EMRA_, CD45RA^+^ CCR7^-^) are indicated. (C) Graphs showing differences in frequencies of CD8 (left) and CD4 (right) T cells specific for S_Delta_ and S_Omicron_ in patients before (left) and 30 days after (right) the third vaccine dose. Wilcoxon signed ranks test was used.

## Notes

### Author Declarations

This prospective observational study has been carried out since March 2021 at the Geneva University Hospitals, Switzerland, according to the principles of Good Clinical Practice and was approved by the Geneva Cantonal Ethics Commission (2021-00430)

