## Supplementary figures and images for "Omicron-specific cytotoxic T-cell responses are boosted following a third dose of mRNA COVID-19 vaccine in anti-CD20-treated multiple sclerosis patients"

### supplementray fig 1

#### Supplemental Figure 1

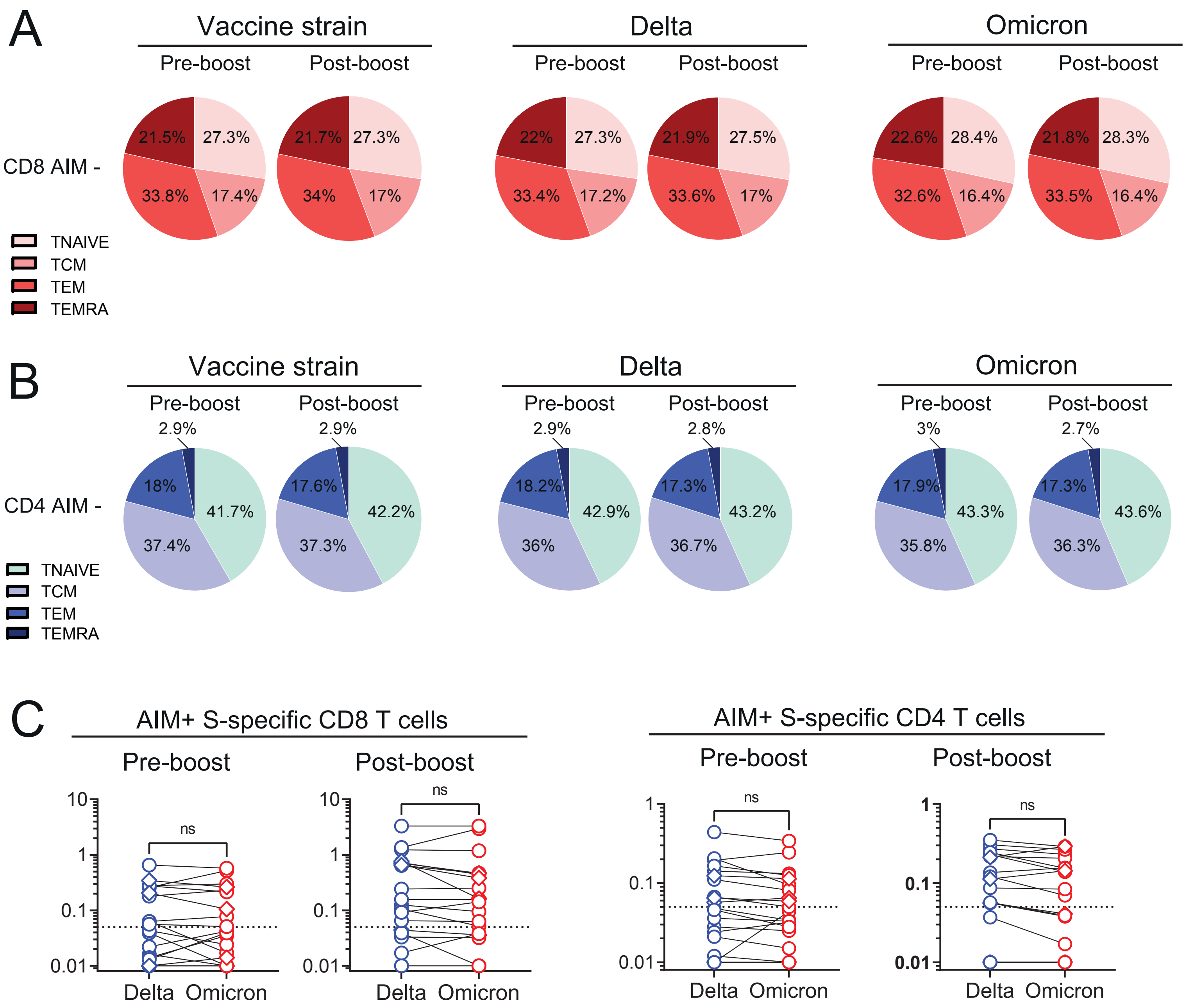
